# Outpatient and Home Pulmonary Rehabilitation Program Post COVID-19: A study protocol for clinical trial

**DOI:** 10.1101/2022.04.08.22273608

**Authors:** Luis V. F. Oliveira, Miriã C. Oliveira, Maria E. M. Lino, Marilucia M. Carrijo, João Pedro R. Afonso, Ricardo S. Moura, Adriano L. Fonseca, Daniela R. P. Fonseca, Luis Felipe R. J. Oliveira, Letícia S. Galvão, Bianca M. Reis, Raphael H. C. O. Diniz, Rubens R. Bernardes, Elisângela R. P. Póvoa, Anderson S. Silva, Dante B. Santos, Vinicius Z Maldaner, Jean Carlos Coutinho, Guilherme Pacheco Modesto, Iransé Oliveira-Silva, Rodrigo A. B. Lopes Martins, Patrícia S. L. Lopes Martins, Claudia S. Oliveira, Gerson Cipriano Júnior, Rodolfo P Vieira, Renata K. Palma, Larissa R. Alves, Giuseppe Insalaco

## Abstract

**Background:** The coronavirus disease 2019 (COVID-19) is a widespread, highly contagious inflammatory process that causes respiratory, physical and psychological dysfunction. COVID-19 mainly affects the respiratory system and evolves in the acute phase from mild cases with common symptoms, such as fever, cough, and fatigue, to the moderate-to-severe form, causing massive alveolar damage resulting in dyspnea and hypoxemia that can rapidly progress to pneumonia, and acute respiratory distress syndrome. The acute form usually causes severe pulmonary sequelae such as pulmonary fibrosis or progression to organ failure, leading to worsening metabolic dysfunction and/or death.

**Purpose:** To verify the effects of an outpatient and home pulmonary rehabilitation program (PRP) on clinical symptoms, pulmonary function, physical activity level, functional status, autonomic activity, peripheral muscle strength, static and functional balance, functional mobility, anxiety and depression, post-traumatic stress, health-related quality of life, and survival of patients with sequelae from COVID-19.

**Methods:** This study will be a cohort, parallel, two-arm multicentric study, to be carried out in three clinical centers, with blind evaluation, with 06 weeks of training and follow-up. This study was designed according to the recommendations of the CONSORT statement. To be involved in this clinical study, according to the inclusion criteria, women and men aged between 16 and 75 years affected by COVID-19. The proposed PRP is based on the guidelines recommended by the Global Initiative for Chronic Obstructive Lung Disease and, consists of a combination of aerobic and muscle strengthening exercises, lasting six weeks, with a frequency of three times a week.

**Discussion:** In patients infected with COVID-19 with persistent symptoms and sequelae, PRP mainly seeks to improve dyspnea, relieve anxiety and depression, prevent, and reduce complications and/or dysfunctions, reduce morbidity and mortality, and improve health-related quality of life.

**Trial registration:** This study was registered at clinicaltrials.gov (ID: COVID-19 PULMONARY REHAB NCT04982042).

## Introduction

The disease caused by coronavirus disease 2019 (COVID-19) is characterized by a widespread, highly contagious inflammatory process that causes respiratory, physical, and psychological dysfunction in patients. COVID19 emerged in December 2019 in Wuhan, Hubei province, China, and alarmingly spread worldwide, with the growth in the number of cases and deaths being considered a global outbreak with a dramatic impact. It was declared a pandemic by the World Health Organization (WHO) in March 2020 [1].

Severe acute respiratory syndrome coronavirus 2 (SARS-CoV-2), caused by COVID-19, mainly affects the respiratory system and evolves in the acute phase from mild cases with common symptoms, such as fever, cough, and fatigue, to the moderate-to-severe form, causing massive alveolar damage resulting in dyspnea and hypoxemia that can rapidly progress to pneumonia, acute respiratory distress syndrome (ARDS), difficult-to-correct metabolic acidosis, and clotting disorders. The acute form of COVID-19 usually causes severe pulmonary sequelae such as pulmonary fibrosis or progression to multiple organ failure, leading to worsening metabolic dysfunction and/or death [2–4]. Hospital admission rates for COVID-19 patients have been difficult to estimate because they depend on the prevalence of community testing and admission criteria, which vary between countries [5]. The WHO reported that patients with COVID-19 develop signs and symptoms after 5 to 6 days of infection, although asymptomatic cases range from 27% to 40% of cases, as indicated by pre-vaccination data [6]. The mean time interval from the onset of the first symptoms of dyspnea to hospitalization and ARDS has been 5 to 8 days, respectively, and in this same period, the scientific literature estimates that 80% of cases were asymptomatic or mild, 15% severe, and 5% critical [7–9]. Elderly, immunosuppressed, and hypertensive people with underlying chronic heart or lung disease are at a greater risk of developing the severe form of COVID-19, requiring admission to an intensive care unit (ICU) and progressing to a worse prognosis [10,11].

In view of the broad-spectrum clinical manifestations associated with multi-organ involvement in patients infected with SARS-CoV-2, the main acute complications are scientifically well-delineated; however, late outcomes are still in the recognition phase [12]. Considering the potential medium- and long-term sequelae in COVID-19 survivors, several organizations, including the WHO and the International Severe Acute Respiratory and Emerging Infection Consortium (ISARIC) have joined forces in scientific protocols and in the development of international recommendations to identify and define the severity of persistent manifestations after ICU and/or outpatient hospitalization [13].

The most common complaints reported in scientific literature after COVID-19 are caused by physiological, immunological, and inflammatory changes in response to infection, either directly by the virus or indirectly by the patient’s immune response [14,15]. The main complaints were dyspnea and persistent fatigue followed by acquired muscle weakness, pulmonary dysfunction, intolerance to moderate efforts, myalgia and arthralgia, post-traumatic stress disorder (PTSD), anxiety, and depression, which justify the worsening of functional status and quality of life in these patients [11,12,16,17]. The fact that the frequency and severity of post-disease symptoms are not directly associated or are exclusive to patients in the risk group or who develop the severe form makes it an urgent need to characterize risk predictors and the potential differences between post-intensive care syndrome and the assignment of rehabilitation programs to minimize sequelae [18,19].

The scientific community and health professionals have discussed the challenge of rehabilitating these patients who present with an infinity of clinical manifestations, considering that knowledge is an important variable in formulating guidelines aimed at structuring the organization, clinical treatment, and pulmonary rehabilitation of these patients with a focus on the implementation of preventive and therapeutic measures [20–22]. The American Thoracic Society and European Respiratory Society (ATS/ERS) task force advocates the inclusion of COVID-19 survivors in a pulmonary rehabilitation program (PRP) lasting 6–8 weeks. The initial experience in China also indicates that a 6-week program can improve lung function, quality of life variables, anxiety, and depression [23,24].

Pulmonary rehabilitation is defined as “a multidisciplinary intervention based on personalized assessment and treatment, which includes exercise training, education, and behavioral modification designed to improve the physical and psychological condition of people with respiratory disease” [25]. In patients infected with COVID-19 with persistent symptoms and sequelae, pulmonary rehabilitation mainly seeks to improve dyspnea, relieve anxiety and depression, prevent and reduce complications and/or dysfunctions, reduce morbidity and mortality, and improve health-related quality of life as much as possible [26,27].

Aerobic exercises, training of the ventilatory muscles, and muscle strengthening of the lower and upper limbs should be progressively offered to patients so that they can gradually recover the level of physical activity observed before the onset of the disease and eventually return to normal activities of daily living (ADLs). The principle of personalization must be respected regardless of the type of intervention in a PRP, based on the specific dysfunction of each patient [28–30].

## Objectives

To verify through a clinical, prospective, and consecutive study the effects of an outpatient and home pulmonary rehabilitation program on clinical symptoms, pulmonary function, physical activity level, functional status, autonomic activity, peripheral muscle strength, static and functional balance assessment, functional mobility, anxiety and depression, post-traumatic stress, quality of life, and survival of patients with pulmonary sequelae from COVID-19.

## Materials and Methods

### Study design

This study will be a cohort, parallel, two-arm multicentric study, to be carried out in three clinical centers, with blind transversal and longitudinal evaluation, with 06 weeks of training and follow-up. This clinical trial was designed according to the recommendations of the CONSORT statement [31]. This study protocol has been designed in accordance with the SPIRIT 2013 Guidelines [32]. Currently, this clinical trial is recruiting patients. The schedule of enrollment, interventions, and assessments according to the SPIRIT guidelines are shown in Figure 1. All procedures of the study adhere to the CONSORT guidelines and are summarized in a flowchart (Figure 2).

**Figure 1.**
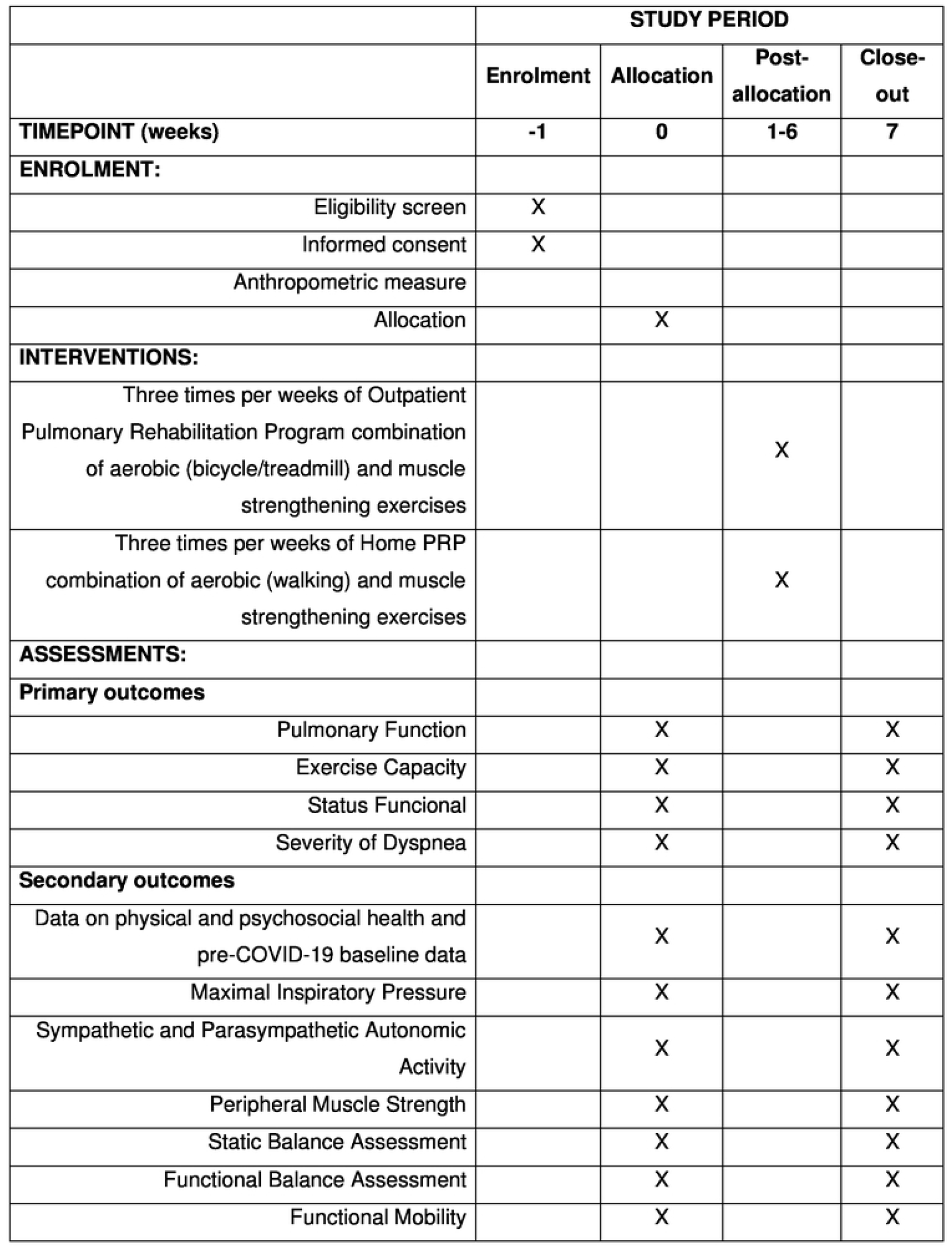
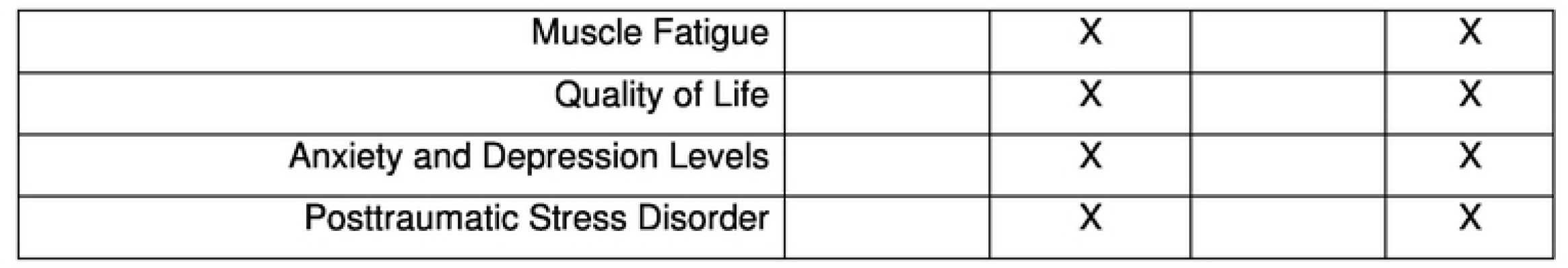
Schedule of enrollment, interventions, and assessments from SPIRIT guidelines.

**Figure 2.**
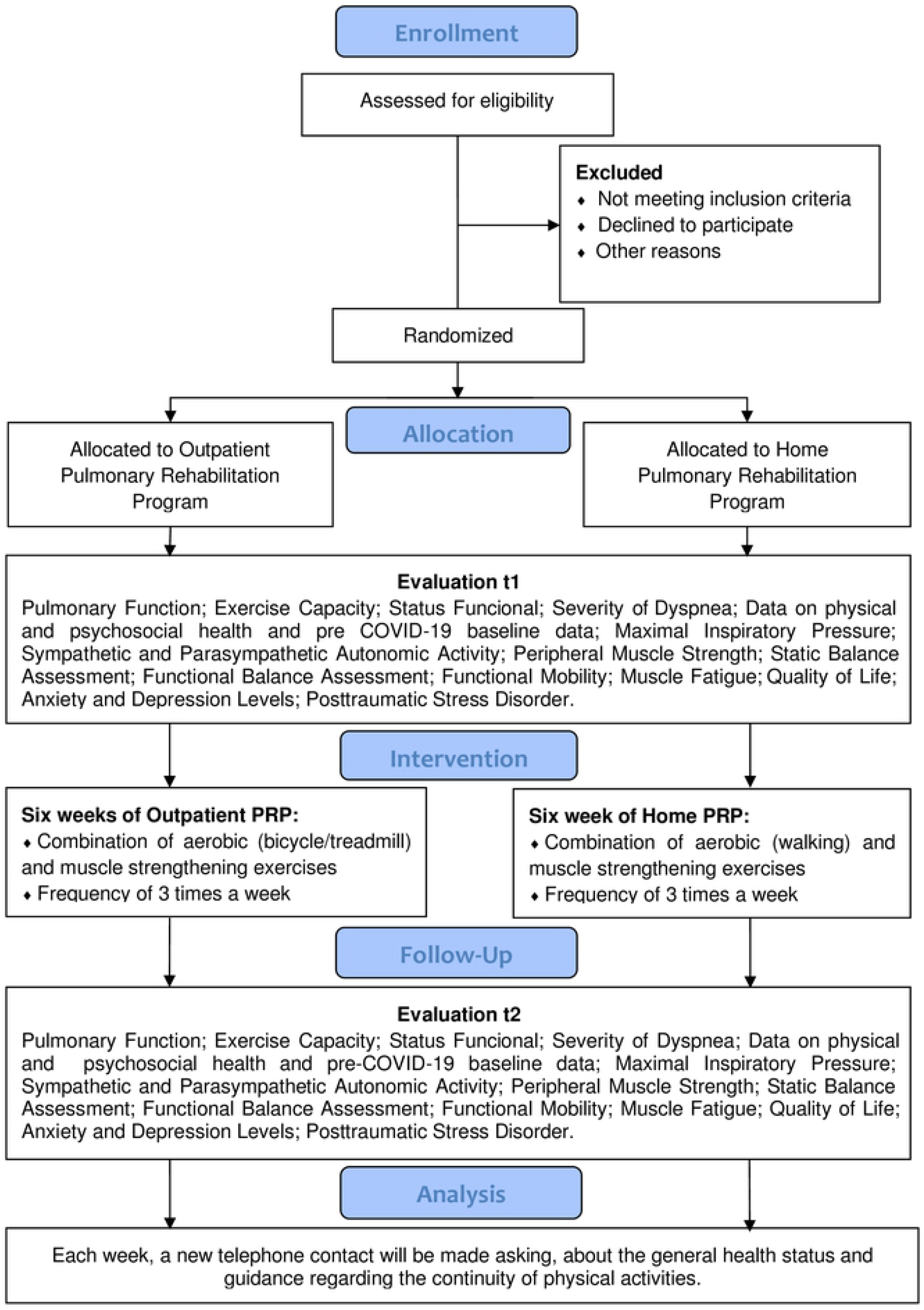
Study flowchart in accordance with the CONSORT statement.

### Study setting

The study will be conducted at the Pulmonary Rehabilitation Laboratory of the Evangelical University of Goiás, UniEVANGÉLICA. Recruitment will take place between July 2021 and December 2022 through social media and banners distributed in reference hospitals for the treatment of patients with COVID-19 in the city of Anapolis (GO), Brazil. The city of Anápolis, in the interior of the state of Goiás, in the Midwest region of Brazil, is considered a center of reference in health in the region, receiving patients from several surrounding municipalities. Many patients who seek care in the city come from neighboring cities and therefore do not have an assistance service in the field of pulmonary rehabilitation, and international biosafety standards for protection against COVID-19 will be observed.

### Ethical considerations and consent to participate

All procedures performed with the patients involved in this study were in accordance with the international ethical standards for research involving human. The research protocol was approved by the Ethics Committee for Research with Human Beings of Universidade Evangélica de Goiás (UNIEVANGÉLICA) on September 24, 2020, under protocol number 4,235,203. This clinical trial was registered at ClinicalTrials.gov (ID: COVID-19 PULMONARY REHAB NCT04982042). Involvement in the study will be voluntary, and leave will be allowed at any time without any burden. Participants will be required to sign an informed consent form after receiving all information about the pulmonary rehabilitation program.

### Participants: recruitment and eligibility criteria

To be involved in this clinical study, according to the inclusion criteria, women and men aged between 16 and 75 years affected by COVID-19 will be invited.

### Inclusion criteria

The following are the inclusion criteria:

Age 16–75 years

Patients who have been affected by COVID-19 and with impaired lung function

Clinically stabilized patients

Patients who agreed to participate in the pulmonary rehabilitation program for at least 12 weeks and signed a free and informed consent form

### Exclusion criteria

The following are the exclusion criteria:

Hospitalized patients

Patients affected with chronic neurological diseases that make it impossible to understand and perform physical activities

Patients suffering from hypertension and cardiovascular conditions without medical treatment

Patients affected with acute phase of rheumatologic disorders

Patients who have recent musculoskeletal disorders and who are not fully recovered from their injuries

Patients affected with chronic mental and/or psychological disturbances

Presence of terminal neoplastic disease

Red flags for serious conditions (night pain, severe muscle spasm, loss of involuntary weight, and symptom mismatch)

Patients classified as severe cases according to the following criteria will also be excluded.

- Respiratory rate ≥ 30 rpm
- SpO2 < 90%
- Cardiac rate > 125 bpm
- Hypotension (systolic blood pressure < 90 mmHg or diastolic blood pressure < 60 mmHg)
- Severe dyspnea (minimal effort or rest)
- Hemoptysis
- Altered alertness (lethargy, acute confusion, disorientation)
- Inability to consume orally due to unintended vomiting or a significant number of bowel movements (≥ 10 per day), suggesting dehydration or hydroelectric disturbances
- Significant impact on general health condition
- A high clinical suspicion of pneumonia requiring radiography, based on worsening of dyspnea, more than 7 days with fever, respiratory rate higher than 22 rpm, and alteration in auscultation findings.

### Interventions

Initially, all patients who met the inclusion criteria underwent clinical, physical, and psychological evaluations. They will then perform pulmonary function tests [33], peripheral muscle strength tests [34], and the 6-min walk test (6 MWT), in accordance with the standards recommended by the ATS [35]. The modified dyspnea scale of the Modified Medical Research Council [36] and the scales for anxiety and depression [37] will also be applied.

The proposed outpatient PRP is based on the guidelines recommended by the Brazilian Society of Pulmonology [38] and by the Global Initiative for Chronic Obstructive Lung Disease [39]. The Outpatient Pulmonary Rehabilitation Program consists of a combination of aerobic and muscle strengthening exercises, lasting six weeks, with a frequency of three times a week, every morning, according Table1. The pulmonary rehabilitation program consists of warm-up, training, and cool-down phases. The warm-up phase consisted of intercalated calisthenic exercises for the muscle groups of the lower and upper limbs according to each patient’s tolerance. The aerobic training phase is performed on a treadmill or bicycle according to the patient’s preference. During the cooling-down phase, muscle stretching, and relaxation exercises are performed in a calm and silent environment.

**Table 1.** Combined aerobic exercise and muscle strengthening exercises training periodization.

|  | **Outpatient PRP** |  | **Home PRP** |  |
| --- | --- | --- | --- | --- |
|  | **Week 1-3** | **Week 4-6** | **Week 1-3** | **Week 4-6** |
|  | Sets x<br>Repetitions | Sets x<br>Repetitions | Sets x<br>Repetitions | Sets x<br>Repetitions |
| **Aerobic exercise** |  |  |  |  |
| Bicycle/treadmill or walking |  |  |  |  |
| Volume | 30 minutes | 40 minutes | 30 minutes | 40 minutes |
| Intensity, % HR 6MWT | 60-70%<br>(4-5 RPE) | 70-80%<br>(5-6 RPE) | 50-60%<br>(4-5 RPE) | 60-70%<br>(5-6 RPE) |
| **Muscle strengthening exercises** |  |  |  |  |
| Upper limb |  |  |  |  |
| Upper limb diagonal (D1, adapted kabat) | 3 x 10-12 | 3 x 13-15 | 3 x 10-12 | 3 x 13-15 |
| Upper limb diagonal (D2, adapted kabat) | 3 x 10-12 | 3 x 13-15 | 3 x 10-12 | 3 x 13-15 |
| Lower limb |  |  |  |  |
| Hip flexors with extended knee | 3 x 10-12 | 3 x 13-15 | 3 x 10-12 | 3 x 13-15 |
| knee extenders | 3 x 10-12 | 3 x 13-15 | 3 x 10-12 | 3 x 13-15 |
| knee flexors | 3 x 10-12 | 3 x 13-15 | 3 x 10-12 | 3 x 13-15 |
| plantar flexors | 3 x 10-12 | 3 x 13-15 | 3 x 10-12 | 3 x 13-15 |
| Volume | 20 minutes | 30 minutes | 20 minutes | 30 minutes |
| Intensity, % 1RM | 50-60%<br>(4-5 RPE) | 60-70%<br>(5-6 RPE) | 50-60%<br>(4-5 RPE) | 60-70%<br>(5-6 RPE) |

PRP: Pulmonary Rehabilitation Program; HR heart rate; 6MWT: Six Minute Walk Test; RPE: rating of perceived exertion; 1RM: one repetition maximum test;

### Home PRP

Home PRP will consist of the same combination of warm-up, strengthening, training, and relaxation exercises as the outpatient PRP. Initially, patients in this group will receive exercise program training by a specialized healthcare professional on the research team and will be encouraged to follow the protocol at home. The aerobic conditioning of this group will be performed by walking on a flat terrain for up to 20 min, with an intensity of 60% to 80% of the maximum heart rate reached in the 6 MWT, respecting individual tolerance and self-monitoring throughout the training. You will be asked to complete a diary for each training session. During the 6 weeks of training, individuals will receive weekly phone calls from the same researcher on the team to monitor the load increase, detect any type of problem, clarify possible doubts, and reinforce the importance of rehabilitation. Patients will be recommended to decrease the intensity or discontinue the exercise in case of a high degree of dyspnea or any other symptom of discomfort. The criteria for load increment in the upper- and lower-limb exercises will be the same as those for the outpatient group.

Upon completing 6 weeks of activities in the pulmonary rehabilitation program, patients will be recruited for a new evaluation, discharged, and encouraged to continue physical activities inherent to PRP at home. Each month, a new telephone contact will be made by a blinded evaluator asking about the general health status, adverse effects, and guidance regarding the continuity of physical activities.

### Outcomes and measurements

All data regarding the initial and final evaluations after 6 weeks of training will be collected by two physiotherapist researchers from the team. The collected data will be registered in standard forms for each outcome and then entered into an Excel database for further analysis. Pulmonary function tests will be performed by a technician trained in pulmonary function and interpreted by a pulmonologist. Physical therapists will perform muscle strength and exercise capacity tests. Quality of life questionnaires and anxiety, depression, and stress scales will be administered by an expert psychologist. All physical activities in the PRP will be monitored by two physiotherapists, who will continuously check the patients’ vital signs. Below, we present the outcomes that will be evaluated in the baseline evaluation and after 6 weeks of participation in the program. The outcomes, measures, and assessment methods adopted in this clinical trial are shown in Table 2.

**Table 2.** Outcomes, measures, and assessment methods that will be used in this clinical trial.

| OUTCOMES | MEASURES | ASSESSMENT |  |
| --- | --- | --- | --- |
|  |  | t1 | t2 |
| Pulmonary Function (FVC, VEF<sub>1</sub>, | Spirometry test | X | X |
| VEF<sub>1</sub>/ FVC) |  |  |  |
| Exercise Capacity | 6MWT | X | X |
| Status Funcional | Post-COVID-19 Functional Status Scale | X | X |
| Severity of Dyspnea | MRC Dyspnea Scale | X | X |
| Data on physical and, psychosocial health and pre-COVID-19 baseline data | ISARIC Global Tier 1 COVID-19 Follow Up Survey. v1.2 21 Jan. 2021 | X | X |
| Maximal Inspiratory Pressure | POWERbreath | X | X |
| Sympathetic and Parasympathetic Autonomic Activity | Heart Rate Variability Analyses | X | X |
| Peripheral Muscle Strength | Handgrip dynamometer | X | X |
|  | MRC-SS | X | X |
| Static Balance Assessment | Force Platform | X | X |
| Functional Balance Assessment | Berg Balance Scale | X | X |
| Functional Mobility | TUG test | X | X |
| Muscle Fatigue | FSS | X | X |
| Quality of Life | SF-36 Questionnaire | X | X |
|  | EQ-5D-5L | X | X |
| Anxiety and Depression Levels | HADS | X | X |
|  | SAS/SDS | X | X |
| Posttraumatic Stress Disorder | IES-6 | X | X |

t1: before intervention; t2: after intervention; FVC: Forced vital capacity; VEF<sub>1</sub>: Forced expiratory volume in one second; 6MWT: Six Minute Walk Test; mMRC: modified Medical Research Council; ISARIC: International Severe Acute Respiratory and emerging Infection Consortium; MRC-SS: Medical Research Council Sum-Score; TUG: Timed Up and Go; FSS: Fatigue Severity Scale; SF- 36: Short Form-36; EQ-5D-5L: EuroQol five dimension five levels; HADS: Hospital Anxiety and Depression Scale; SAS/SDS: The Self-Assessment of Anxiety Scale and the Self-Assessment of Depression Scale; IES-6: Event Impact Scale - 6.

### Primary outcome

#### Pulmonary function: spirometry test

The Koko Sx 1000 spirometer (Koko PFT, Fordham, Longmont, CO, USA) will be used according the guidelines of the ERS, ATS [40, 41], and Brazilian Society of Pneumology [42]. Spirometry tests will be performed on all patients in the morning after two puffs of salbutamol (400 μg), which will be administered using a spacer (Volumatic, Glaxo Smith Kline LTD, London, UK). The best forced expiratory volume in one second (FEV1) and forced vital capacity (FVC) were chosen for all analyses, regardless of the best curve. The ATS/ERS acceptability criteria for spirometry will be applied, including a minimum of three respiratory maneuvers with at least two being free from artifacts.

#### Exercise capacity: walk test (6 MWT)

According to the guidelines published by the ATS (2002), the 6 MWT evaluates the distance a patient can walk quickly for a maximum period of 6 MWD. This test assesses the global and integrated responses of all systems involved in physical activities. [43] The 6 MWT is a simple, reliable, practical, low-cost, safe, and easy-to-apply functional capacity measure that can be performed in a flat, rigid space of 30 m. [44–46]

The test has been widely used to assess the functional status of patients with cardiorespiratory impairment and is highly correlated with morbidity and mortality [47–51]. An interesting review concluded that the 6 MWT is easy to administer, better tolerated, and more representative of ADLs than other walking tests [52].

#### Status Functional status: Post COVID-19 Functional Status Scale (PCFS)

Due to the wide variety of clinical symptoms and sequelae presented by patients after COVID-19, it is of great importance to have a simple and reproducible tool to assess the impact of the disease, monitor its evolution, and guide rehabilitation actions [53,54]. Given the high incidence of pulmonary embolism with myocardial damage/myocarditis and neurological complications in patients with critical-stage COVID-19, Dr. Klok et al. proposed the use of the PVFS scale (with a small adaptation) in the evaluation of patients after COVID-19. The authors suggest the use of the PVFS scale at hospital discharge, 4 and 8 weeks after discharge, and 6 months to assess the recovery of these patients. They also recognized that this instrument should not replace other established and validated instruments in the assessment of quality of life, tiredness, or dyspnea but could also be used as a tool for evaluating the results of the functional state of intervention actions after COVID-19 [55,56].

This instrument covers functional level, limitations in daily life tasks, and lifestyle changes at six levels. Level 0 demonstrates the absence of any functional limitation, and the death of a patient is recorded in grade D. From level 1 onwards, symptoms, pain, or anxiety are presented in ascending order. This had no effect on the activities of patients in grade 1, whereas a lower intensity of activities was required for those in grade 2. Level 3 is characterized by the inability to perform certain activities, forcing patients to adapt. Finally, level 4 is reserved for patients with severe functional limitations who need assistance in performing ADLs [57, 58].

#### Severity of dyspnea: Medical Research Council (MRC) Dyspnea Scale

In patients with chronic obstructive pulmonary disease (COPD), it has been observed that the interaction between dyspnea, physical deconditioning, and muscle weakness results in a vicious circle or negative spiral [59], which generates important functional limitations [60]. In real life, these functional limitations are characterized by a reduction in the ability to perform ADLs [61]. It has been observed that a close relationship exists between physical ADL, morbidity, and mortality in patients with COPD, calling attention to the adequate evaluation and follow-up of their evolution [62–64]. In this study, owing to its ease of application and understanding, the MRC scale will be used, an instrument traditionally used in the international literature [65,66].

The MRC scale consists of five items, where the patient defines the item that best corresponds to the extent to which dyspnea limits daily activities. The patient reported his subjective degree of dyspnea, choosing a value between 1 (shortness of breath only during intense exercise) and 5 (shortness of breath when getting dressed or feeling short of breath that he no longer leaves the house) [67,68].

### Secondary outcome

#### Physical and psychosocial health COVID-19: International Severe Acute Respiratory and Emerging Infections Consortium (ISARIC)

ISARIC has recently published the COVID-19 response timeline to mark the amazing work performed by all those using ISARIC’s resources and clinical data platform since the beginning of the pandemic; the timeline showcases how the federation has taken action right from the start of the pandemic to address the most important research questions to further the knowledge of COVID-19, thereby saving lives and providing a collaborative platform through which global, patient-oriented clinical studies can be developed, executed, and shared.

Protocols addressing the most important questions between and during epidemics of severe acute respiratory infections and other rapidly emerging public health threats are undertaken to generate new knowledge, maximize the availability of clinical information, and thereby save lives.

The ISARIC COVID-19 follow-up protocol was based on the ISARIC/WHO COVID-19 Clinical Characterization Protocol. It assesses the risk and risk factors for long-term physical and psychosocial health consequences following COVID-19 diagnosis using a range of validated tools. This protocol will follow-up patients with confirmed COVID-19 using standardized data collection forms. The forms can be completed as patient self-assessment via post or an online link, or via clinician/research-led completion via telephone or in-clinic. It can be used to identify people for further in-clinic follow-up and assessment or in conjunction with sampling and diagnostic studies.

Assessment of the risk factors for long-term sequelae requires data on pre-existing conditions and care received during the acute phase of the disease. In this way, ISARIC created tools for researchers to collect and store clinical data in a standardized manner through COVID-19 follow-up surveys. The Tier 1 Initial Follow-up Survey will be used in the initial assessment of patients, and the Tier 1 Ongoing Follow-up Survey will be used in the reassessment after six weeks of intervention. The original versions were made available in English and were later translated into other languages, including Portuguese [69,70].

This is an open-access tool that collects data on physical and psychosocial health, occupational status, and socioeconomic variables, as well as pre-COVID-19 baseline data that make it possible to characterize the physical and psychosocial repercussions after hospital discharge and/or sharp. These include vaccination data, hospitalization and possible decompensations and/or readmissions, specific consequences including deep vein thrombosis (DVT), stroke or transient ischemic attack, pulmonary embolism, recent febrile illness, new or persistent symptoms, quality of life (assessed using the EQ-5D-5L and EQ VAS), dyspnea (measured by the MRC dyspnea scale), difficulties in ADLs due to health problems (UN/Washington disability score), and lifestyle [71].

#### Maximal inspiratory pressure (MIP): POWERbreath

The pressure generated by the inspiratory muscles will be quantified using an electronic threshold device (PowerBreathe Medic KH2, IMT Technologies Ltd., Birmingham, UK). It features an advanced live feedback software (Breathe-Link Medic), which allows patient results to be viewed in real time. To measure the MIP, patients should be comfortably seated, encouraged to perform a maximal expiratory effort from the residual volume, and then a verbal command will be given to perform a maximal inspiratory effort, according to ATS/ERS standards [72]. The reference values used were described by Pessoa et al. (2014), based on parameters for the Brazilian population [73].

#### Nervous autonomic activity: heart rate variability (HRV) analyses

HRV was analyzed using Kubios HRV software (Kubios Ou Limited company, Kuopio, Finland). This software is widely known and is used worldwide with different patient populations and healthy individuals. It is a well-developed, validated, and easy-to-use software for analyzing the behavior of sympathetic and parasympathetic autonomic activities. This software is considered to be the gold standard for HRV analysis. In this clinical research protocol, HRV analysis will be performed during the 6 MWT. Data will be collected using the Kubios software from the heart rate and time, obtaining the time values referring to the R-R intervals. Five minutes will be collected with the patient seated, at rest before starting the test, during the 6 min of walking and for 5 min after the test, and sitting at rest. HRV analyses will be performed in the time and frequency domains, as well as in nonlinear models.

### Peripheral muscle strength

#### Handgrip dynamometer

According to the recommendations of the American Society of Hand Therapists, handgrip strength should be assessed using a dynamometer on both sides (dominant and non-dominant). In this protocol, hand grip strength will be verified using the digital hand dynamometer Jamar® Plus Hand Dynamometer (JAMAR® Hydraulic Hand Dynamometer, Sammons Preston, Bolingbrook, IL, USA). Patients should be seated with their feet firmly on the floor, shoulders adducted, elbows flexed at 90º, and forearms in a neutral position [74], and the position of the equipment handle should be chosen by the patient himself [75]. Three tests will be performed on each side, alternately, with a rest period of at least 1 min between attempts on the same hand [76]. The highest value observed in each upper limb was used to represent maximum handgrip strength [77]. The dynamometer was adjusted according to the size of the hand of each patient. After instruction, the participant pressed the dynamometer until maximum strength was displayed, and the results were recorded in kilograms to one decimal place [78].

#### MRC sum-score (MRC-SS)

The MRC-SS assesses the global peripheral muscle strength of six upper and lower limb muscle groups (shoulder abduction, elbow flexion, wrist extension, hip flexion, knee extension, and ankle dorsiflexion) on both sides, from 5 (normal strength) to 0 (no visible contraction). The sum of the scores ranges from 0 to 60, where scores below 48 indicate significant peripheral muscle weakness and scores below 36 indicate severe weakness. The therapist’s hand placement and patient positioning were standardized [79].

Muscle strength will initially be evaluated against gravity, and the movements to be evaluated will be passively demonstrated. If the patient could not perform the movement, positioning was modified [80]. An isometric resistance was applied to the end of the range of motion to test each degree of muscle strength. This score has been widely used for the diagnosis of ICU-acquired weakness (ICU-AW) [81,82].

#### Functional balance assessment: Berg balance scale

The Berg balance scale assesses functional balance by performing 14 tasks of daily living, with a global score of 56 possible points. Items were scored from 0-4, with 0 representing the inability to complete the task and 4 representing the ability to complete the proposed task. A score from 0 to 20 represents impairment of balance, 21 to 40 represents acceptable balance, and 41 to 56 represents good balance [83].

#### Functional mobility: timed Up & Go (TUG)

The TUG test is an instrument that allows the assessment of functional mobility in different populations as a function of time [84,85]. To perform the test, the patient had to get up from a standardized chair, walk 3 m at a comfortable pace, return to the chair, and sit down. The test will be performed with the subjects on barefoot/shoes three times, the first time for familiarization. Time intervals above 16 s are indicate impairment of the patient’s functional mobility and identified as a high fall risk [86].

TUG test data were obtained using a wireless inertial detection device: Inertial Sensor: G-Sensor® (BTS Bioengineering Corp., Quincy, MA, USA). A portable Gsensor is a wireless inertial sensor system used to analyze human movements. The sensors were controlled by a data logging unit (up to 16 elements) via ZigBee-type radio communication. Each sensor has dimensions of 62 mm × 36 mm × 16 mm, weighs 60 g, and is composed of a three-axis accelerometer (maximum scale ± 6 g), a 3-axis gyroscope (full scale ± 300°/s), and a magnetometer 3-axis (full scale ± 6 Gauss). This device was calibrated with gravitational acceleration immediately after manufacturing. For this study, only one device was used to collect data at a sampling frequency of 50 Hz. The data from the inertial sensor were transmitted via Bluetooth to a computer and processed using proprietary software (BTS GSTUDIO, version: 2.6.12.0), which automatically provided the parameters. The TUG test has been validated in a population of patients with COPD.

#### Static balance assessment: Force platform

Postural balance is assessed using stabilometry, also called oscillometry, which measures oscillations in the standing posture. This condition is measured through the displacement of the center of pressure (COP) while the individual remains standing on the platform for a certain time [87]. It allows detection of the slightest variations in amplitude and frequency of displacement of the body’s center of gravity, helping to analyze the functional aspects related to body imbalance. It is a technique used to evaluate the plantar pressure exerted by the body, which allows the detection of postural oscillations and balance deficits [88].

To assess static balance, the SMART-D 140® System (BTS Bioengineering Corp., Quincy, MA, USA) containing two force platforms (Kistler Platform model 9286BA) was used. The platform acquisition frequency was 100 Hz, captured by four piezoelectric sensors positioned at the extremities of the force platform measuring 400/600 mm. Data were recorded and interpreted using SWAY BTS 161 Engineering software (BTS Bioengineering Corp., Quincy, MA, USA), integrated, and synchronized with the SMART-D 140® system (BTS Bioengineering Corp., Quincy, MA, USA). The collection protocol to be conducted includes the instruction to remain in a standing position, as still as possible, with the arms close to the body and the head kept in a vertical position. Measurements (30 s) of the COP displacement on the X (anterior-posterior), Y (medium-lateral), and COP-GOG axes will be taken under the conditions of eyes open and eyes closed, with the proprioceptive perturbation that configures as a soft surface collecting with open eyes and eyes closed.

#### Muscle fatigue: Fatigue Severity Scale (FSS)

The FSS is one of the most widely used fatigue self-assessment scales worldwide. This scale classifies the severity of a patient’s fatigue symptoms in terms of how they compromise their motivation, physical activity, and ADLs. The scale is composed of a self-report questionnaire with nine items, with scores ranging from 1 to 7, where 1 indicates strongly disagree and 7 indicates strongly agree [89,90]. The scale is scored by the total sum or by the calculation of an average score on the nine items, with higher scores indicating more severe fatigue. A score ≥ 4 indicated fatigue [91]. The FSS has been translated and validated in several languages including Portuguese. Participants will be asked to respond to the FSS relative to the previous week [92].

### Health-related Quality of life

#### Short Form Health Survey 36 (SF-36) questionnaire

In this protocol, health-related quality of life is assessed using a generic instrument. The SF-36 is a multidimensional questionnaire frequently used to assess the quality of life of patients. This instrument was developed by Ware and Sherbourne [93] and validated in Brazil by Ciconelli et al. [94]. It consists of 36 questions that analyze eight health domains: functional capacity, limitations due to physical aspects, pain, general health status, vitality, social aspects, limitations due to social aspects, and mental health [95]. A final score ranging from 0 to 100 is generated from each domain, and the higher the score, the better the health-related quality of life [96].

#### EuroQol five dimension five levels (EQ-5D-5L)

The EQ-5D-5L version was introduced by the EuroQol Group in 2009 with the aim of improving the instrument’s reliability and sensitivity compared to the EQ-5D-3L. The EQ-5D-5L consists of the EQ-5D descriptive system, covering five dimensions: mobility, self-care, usual activities, pain/discomfort, and anxiety/depression. Each dimension has five levels of severity: none, mild, moderate, severe, and extreme. The patients will be asked to indicate their health status in each of the five dimensions. A score can be calculated from the scores generated by each domain, with higher scores representing a better quality of life [96–98].

### Anxiety and depression levels

#### The Hospital Anxiety and Depression Scale (HADS)

Several instruments in the scientific literature have been used to assess anxiety and depression [99] The HADS was created 30 years ago by Zigmond and Snaith to identify symptoms of anxiety and depression in non-psychiatric inpatients and later on in outpatients and other types of patients [100]. The HADS has been validated for use in Brazil by [101], and it is short and takes a few minutes to complete while waiting for an appointment with the doctor. The HADS uses a 4-point response scale (0–3) according to the severity of anxiety and depression. The depression domain was measured using seven items, and the other seven items measured the anxiety domain. The total score for each domain ranges from 0 to 21. In both domains, cases with scores from 0 to 7 points were considered normal, cases from 8 to 10 points were considered to have a borderline abnormality, and scores from 11 to 21 were evaluated as cases of anxiety or depression depending on the dotted domain.1.

#### The Self-Assessment of Anxiety Scale (SAS) and the Self-Assessment of Depression Scale (SDS)

The SAS and SDS are instruments commonly used in the assessment of negative emotions in clinical practice and are frequently applied to assess the state of adults with symptoms of anxiety and depression [102].

Each scale is composed of 20 items, using a four-level score formulated as positive (1–4) or negative (4–1). The SAS identifies the presence of affective symptoms based on diagnostic criteria corrected in the American psychiatric literature and assesses four dimensions: cognitive, motor, vegetative, and central nervous system. The total scale score was the sum of the scores for each item. A score below 50 indicates anxiety, a score of 50–60 indicates level anxiety, a score of 61–70 indicates moderate anxiety, and a score above 70 indicates severe anxiety.

The choice of items on the SDS scale was based on studies of an analytical factor of depressive symptoms. The cutoff score was 53, indicating that standard scores greater than 53 induced depression [103]. In Brazil, there has already been validation for the SAS, which has been used with excellent reliability [104]. Both scores range from 20 to 80 (raw score), and then these scores are converted into indices ranging from 25 to 100.

#### Posttraumatic stress disorder: Impact of Event Scale-6 (IES-6)

The IES-6 is an abbreviated version of the Impact of Event Scale-Revised (IES-R), which consists of six five-point items, ranging from 0 to 4 [105]. The IES-6 is an instrument used to identify individuals with significant symptoms of traumatic stress who need a more extensive evaluation to determine the diagnostic criteria for PTSD, without confirmation of a clinical diagnosis [106]. The IES-6 is a reliable and valid tool for screening PTSD and survivors of ARDS and is widely used in hospitalized and discharged patients [107]. This scale has been used in some studies to assess the prevalence of and risk factors for PTSD in patients with COVID19 [108].

#### Safety assessment

All patients will be monitored during all pulmonary rehabilitation program activities. Vital signs, such as temperature, heart rate, partial oxygen saturation, and peripheral blood pressure, will be evaluated at the beginning, during, and at the end of each session. Peripheral oxygen saturation and heart rate were continuously monitored using a G-Tech pulse oximeter (Beijing Choice Electronics Technology Co., Ltd., Beijing, China). Peripheral blood pressures will be measured using a clinical Premium aneroid sphygmomanometer (Wenzhou Instruments Co, China) at the beginning and end of the session or if the patient shows any pressure discomfort.

The perceived exertion by the patients was registered throughout the aerobic training phase using the Borg Dyspnea Scale. Training will always be conducted by an experienced physiotherapist, who is a member of the research team. If the systolic and/or diastolic blood pressure reaches the maximum pre-established limits, the patient will be instructed to reduce the effort and/or stop the activity for a few minutes until the pressure stabilizes and returns to normal values. In view of this clinical situation, the patient was referred to a specialist physician for clinical re-evaluation.

#### Sample size and power calculation

The sample size was calculated according to Shahin et al. (2008). This calculation is based on the mean difference in the distance covered in the test, which should be clinically significant. Considering a difference within +8 (control) and +46 (intervention), for a sample size of 35 and 36 patients, respectively, and an SD within each group of 9.2, an α error probability of 0, 05, and a power (1 - error probability β) of 0.80 for repeated measures, for repeated measures analysis (between factors), a total sample size of 36 individuals was needed to reach a power of 81.77%. An additional 10% of subjects were required to adjust for other factors, such as dropouts, lost data, and loss to follow-up, which resulted in a total of 40 subjects per group. G*Power Statistical Power Analyses for Mac were utilized according to the appropriate reference [109,110].

#### Statistical analyses

The Kolmogorov–Smirnov test was used to test the normality of the distribution of the verified variables. For intragroup comparisons, we will use the Student’s t-test for paired samples that presented parametric distribution or the Mann– Whitney test for variables whose distribution was non-parametric. Intergroup comparisons were made using one-way analysis of variance (ANOVA), and Tukey’s post-test was used for paired comparisons whenever the null hypothesis was rejected by ANOVA. For intergroup comparisons of variables that presented a non-parametric distribution, we used the Kruskal–Wallis test, and the paired comparison (when the null hypothesis was rejected) was made using the Mann–Whitney test. The chi-square test was used to test the association between the groups and exacerbations/hospitalizations. All analyses will be performed using the Statistical Package for Social Science (SPSS) version 23.0 (IBM Corp. Released 2015. IBM SPSS Statistics for Windows, version 23.0. Armonk, NY: IBM Corp). The significance level established for all analyses was set at 5%.

#### Data management

All clinical data of the patients involved in the study will be collected through specific standardized forms for clinical evaluation and stored in a database created by the Scientific Coordination of the Protocol and protected by a password. All patient identification will be replaced by a code to maintain the confidentiality of the data collected. At the end of each activity day, the collected data were backed and exported to another computer. Any changes to the proposed protocol will be immediately communicated to the institution’s Research Ethics Committee and ClinicalTrials.gov.

#### Strategies of study retention

In the initial phase of protocol evaluation, all patients will participate in a health education program at the institution, where they will receive information about the development and progression of the disease, its treatment (both drug and non-drug), the correct use of oxygen, and the importance of participating in a pulmonary rehabilitation program. All patients received a booklet containing the content of the educational program and guidance on the practice of physical activities. During the study period, participants allocated to the home pulmonary rehabilitation group will receive weekly phone calls to monitor the activities performed during the week, check for the presence of adverse events, and encourage and not drop out of the program. Psychological strategies to improve self-esteem will also be adopted by all patients and their caregivers.

## Discussion

International scientific literature has demonstrated the effectiveness of pulmonary rehabilitation in patients with chronic and acute lung diseases. In this sense, we hope that patients affected by COVID-19 undergoing an outpatient and home pulmonary rehabilitation program will obtain benefits in the short-, medium-, and long-term. We highlight the benefits in terms of reduced fatigue and dyspnea, increased functional and exercise capacity, reduced limitations in ADLs, improved quality of life, mood and motivation, increased adherence to recommended clinical treatments, increased participation in therapy decisions, strengthening the patient’s self-management capacity, and reducing the amount of health care for patients, families, and communities, including reducing the number of hospitalizations and increasing survival, with a consequent reduction in health costs for the state.

### Potential impact and significance of the study

According to the international scientific literature, which shows excellent results of pulmonary rehabilitation for patients with pulmonary diseases, we expect that, with the participation of patients affected by COVID-19 in outpatient and home pulmonary rehabilitation programs, we will obtain benefits in the short, medium, and long term. The potential clinical impact of this study will be to reduce fatigue and dyspnea, increase functional and exercise capacity, reduce limitations in ADLs, improve quality of life, and consequently reduce morbidity and mortality in patients affected by COVID-19. A reduction in decompensation and hospitalizations is also expected, representing a reduction in health costs by the government. It is hoped that with the results of this clinical trial, we can encourage the scientific community to increase the availability of pulmonary rehabilitation programs for the large number of patients surviving COVID-19, improve the quality of life, and increase survival.

### Strengths and weakness of the study

Currently, there are a small number of clinical trial protocols for pulmonary rehabilitation programs aimed at patients with post-COVID-19 sequelae according to international clinical trial registry agencies. A strong point of the research is that patients who cannot access the activities of the outpatient pulmonary rehabilitation program will be able to perform the same activities at home. Another strength of this clinical trial is that robust results were not observed in the scientific literature on the clinical response of post-COVID-19 patients undergoing pulmonary rehabilitation programs. One weakness of this clinical trial is the fact that it is not controlled; however, it is justified by the fact that it is unethical to leave patients who present sequelae of COVID-19 without participating in a rehabilitation program.

### Dissemination policy

The results of this clinical trial will be shared with potential users and peers in the field of research, scientific societies, and people formulating public health policies, thus contributing to the progress of science in general and improving the population’s quality of life. The results of this study will be presented at scientific conferences and submitted as a manuscript to scientific journals. The database is made available upon request.

### How amendments to the study, including termination, will be dealt with

Any changes in the execution of the proposed initial protocol and/or adverse events that occur to patients will be communicated to the Research Ethics Committee of the institution, ClinicalTrials.gov, and the health authorities of our country.

## Data Availability

No datasets were generated or analysed during the current study. All relevant data from this study will be made available upon study completion.

https://www4.unievangelica.edu.br/ppg/movimento-humano-e-reabilitacao

## Acknowledgements

The authors would like to thank Fundação de Amparo a Pesquisa do Estado de Goiás (FAPEG) and Universidade Evangélica de Goiás (UniEVANGÉLICA) for their support in all phases of organizing this study protocol.

## Notes

**Funding** This study protocol was approved in an emergency public call from the government of the State of Goiás (Brazil): Public Call: 06/2020 - Mapping of Proposals to Address Covid-19. Administrative process no.: 202010267000277. Research project: “COVID-19 Outpatient and Home Pulmonary Rehabilitation Program” from the Goiás State Research Support Foundation - FAPEG, Brazil. The full name of the funder is: Fundação de Amparo a Pesquisa do Estado de Goiás – FAPEG, Goiania (GO), Brazil. Website: http://www.fapeg.go.gov.br. The authors thank Evangelical University of Goiás - UniEVANGÉLICA for the partial support to the development of this research. The funders had and will not have a role in the study design, data collection and analysis, decision to publish, or preparation of the manuscript. The funding letter has been attached as a supporting file.

### Competing Interest Statement

The authors have declared no competing interest.

### Clinical Trial

ClinicalTrials.gov (ID: COVID-19 PULMONARY REHAB - NCT04982042).

### Funding Statement

The funders had and will not have a role in study design, data collection and analysis, decision to publish, or preparation of the manuscript.

### Author Declarations

All procedures to be performed with the patients involved in this study are in accordance with international ethical standards for research involving human beings as per the 1964 Declaration of Helsinki and its subsequent amendments. This research protocol was approved by the Ethics Committee for Research with Human Beings of the Evangelical University of Goiás (UNIEVANGÉLICA) under protocol number 4.235.203. This clinical trial is registered at ClinicalTrials.gov (ID: COVID-19 PULMONARY REHAB - NCT04982042). Involvement in the study will be voluntary, and leave is allowed at any time without any burden. Participants will be required to sign the Informed Consent Form after receiving all information about the Pulmonary Rehabilitation Program.

